# Who would be affected by a ban on disposable vapes? A population study in Great Britain

**DOI:** 10.1101/2023.10.23.23297396

**Authors:** Sarah E. Jackson, Harry Tattan-Birch, Lion Shahab, Melissa Oldham, Dimitra Kale, Leonie Brose, Jamie Brown

## Abstract

**Objectives:** The UK government is consulting on banning disposable e-cigarettes. This study aimed to describe trends in disposable e-cigarette use among adults in Great Britain since 2021 and establish who would currently be affected by a ban on disposables.

**Study design:** Nationally-representative monthly cross-sectional survey.

**Methods:** We analysed data from 69,973 adults surveyed between January-2021 and August-2023. We estimated monthly time trends in the weighted prevalence of current disposable e-cigarette use among adults and by sociodemographic characteristics and smoking status.

**Results:** From January-2021 to August-2023, the prevalence of disposable e-cigarette use grew from 0.1% to 4.9%. This rise was observed across all population subgroups but was most pronounced among younger adults (e.g., reaching 15.9% of 18-year-olds compared with 1.3% of 65-year-olds), those who currently smoke (16.3%), and those who stopped smoking in the past year (18.2%). Use among never smokers remained relatively rare (1.5%), except among 18-24-year-olds (7.1%). Use was significantly higher in England than Wales or Scotland (5.3% vs. 2.0% and 2.8%) and among less (vs. more) advantaged social grades (6.1% vs. 4.0%), those with (vs. without) children (6.4% vs. 4.4%), and those with (vs. without) a history of mental health conditions (9.3% vs. 3.1%).

**Conclusions:** A ban on disposable e-cigarettes would currently affect one in 20 adults in Great Britain (approximately 2.6 million people). The impact would be greatest on young people, including the 316,000 18-24 year-olds who currently use disposables but who have never regularly smoked tobacco, which may discourage uptake of vaping in this group. However, a ban would also affect 1.1 million people who currently smoke and a further 744,000 who previously smoked. It would also have a disproportionate impact on disadvantaged groups that have higher rates of smoking and typically find it harder to quit.

## Introduction

Early e-cigarettes (‘vapes’) were disposable products that mimicked the look and feel of combustible cigarettes but were poor at delivering nicotine.^1^ Over time, new rechargeable e-cigarette types were developed to deliver nicotine more effectively via refillable tanks or replaceable pods.^2–4^ These devices came to dominate the global e-cigarette market: by 2019, <10% of vapers used disposables.^5–7^ In 2021, a new form of disposable e-cigarette entered the market, and from 2021 to 2022, use of disposables rose sharply in Great Britain as these new products rapidly became popular among young people.^8,9^ Similar trends were observed elsewhere, including among US youth.^10,11^ The UK Government have recently announced a consultation on measures to reduce youth e-cigarette use, including a potential ban on disposable devices.^12^

There are several reasons why new disposable e-cigarettes may appeal to a broader audience than older disposable and reusable devices – including (but not limited to) young people and those who have never smoked. Unlike earlier disposables, these new disposable products are not designed to look like cigarettes, but rather have a sleek design (in a variety of colours) and branding that appeals to young people.^13^ They are promoted through colourful in-store displays, word of mouth, and social media platforms,^14–16^ and are widely available in supermarkets, convenience stores, petrol stations, vape shops, and online without adequate enforcement of age-of-sale laws.^17,18^ They are convenient and easy to use, and have a very low upfront cost, which likely makes them more accessible to experimental users and those with limited money to spend. In addition, they come in various flavours and, like pod devices, deliver nicotine effectively using high-concentration (20mg/ml in EU/UK) nicotine salts e-liquid^19^ which produces an aerosol that may be more palatable for those who have never smoked.^20^

While vaping is much less harmful than smoking, it carries more risks than neither vaping nor smoking.^21^ Preventing uptake of vaping among people who have never smoked, and in particular among young never smokers, is therefore a public health priority. There have been calls for an outright ban on disposable e-cigarettes, given concerns about youth uptake and their environmental impact,^22–24^ and the UK Government is now considering this course of action.^12^ In making decisions about whether or not to ban disposables, it is important for policymakers to understand: (i) how the prevalence of disposable vaping is changing over time within different population groups, (ii) the current size and profile of the population using disposable e-cigarettes in Great Britain who would be affected, and (iii) whether a ban is likely to affect any specific population groups disproportionately. Should the ban be implemented, this knowledge will help to inform ways in which to educate the public about alternative vaping products and plan for the provision of tailored advice and support to vapers (specifically those using disposables).

Using data from a large sample representative of the adult population in Great Britain, collected between January 2021 and August 2023, this study aims to address the following research questions:

1. How has the prevalence of vaping disposables changed since January 2021 among groups with different sociodemographic characteristics and smoking status?
2. As a result of these changes:

a. What is the sociodemographic and smoking profile of those using disposable e-cigarettes in 2023?
b. How does it compare to the profile of those using refillable and pod e-cigarettes in 2023?

## Method

### Pre-registration

The study protocol and analysis plan were pre-registered on Open Science Framework (https://osf.io/uwfd3/). We made one amendment: we had planned to report prevalence ratios for the change in prevalence across the whole time-series among each population group; instead, we report the absolute percentage point change in prevalence from the start to the end of the study period, alongside 95% confidence intervals (CI) calculated using bootstrapping, to better describe differences in the extent of the rise in disposable use across population groups. In addition to our planned analyses, we reported the prevalence of vaping (overall and by device type) among participants surveyed in 2023. We also undertook exploratory analyses to provide more detailed insight into trends in disposable use by age, smoking status, and mental health (described in the statistical analysis section below).

### Design

Data were drawn from the ongoing Smoking Toolkit Study, a monthly cross-sectional survey of a representative sample of adults (≥16 years) in Great Britain.^25,26^ The study uses a hybrid of random probability and simple quota sampling to select a new sample of approximately 2,450 adults each month. Data are collected through telephone interviews. Comparisons with other national surveys and sales data indicate that key variables such as sociodemographic characteristics, smoking prevalence, and cigarette consumption are nationally representative.^25,27^

The present analyses focused on data from respondents in the period from January 2021 to August 2023 (the most recent data available at the time of analysis). Vapers in England were not asked about the type of device they use in May, June, and August 2022, so we excluded adults in England surveyed in these waves to avoid underestimating the prevalence of disposable use. In addition, data on 16- and 17-year-olds were not collected in 2021. These ages were therefore excluded from trend analyses (which used data across the study period) but were included in the comparison of profiles of e-cigarette users in 2023.

### Measures

Vaping status was assessed within several questions asking about use of a range of nicotine products. Current smokers were asked ‘Do you regularly use any of the following in situations when you are not allowed to smoke?’; current smokers and those who had quit in the past year were asked ‘Can I check, are you using any of the following either to help you stop smoking, to help you cut down or for any other reason at all?’; and non-smokers were asked ‘Can I check, are you using any of the following?’. Those who reported using an e-cigarette in response to any of these questions were considered current vapers.

Participants who reported current vaping were asked: ‘Which of the following do you mainly use…?’ Response options were:

a. ‘A disposable e-cigarette or vaping device (non-rechargeable)’
b. ‘An e-cigarette or vaping device that uses replaceable pre-filled cartridges (rechargeable)’
c. ‘An e-cigarette or vaping device with a tank that you refill with liquids (rechargeable)’
d. ‘A modular system that you refill with liquids (you use your own combination of separate devices: batteries, atomizers, etc.)’

Those who responded a were considered current users of disposable e-cigarettes. Those who responded b were considered current users of pod e-cigarettes and those who responded c-d current users of refillable e-cigarettes.

Sociodemographic characteristics included country of residence (England/Scotland/Wales); age; gender (men/women/other – the latter group were excluded from trend analyses by gender due to low numbers); occupational social grade (ABC1 includes managerial, professional and upper supervisory occupations / C2DE includes manual routine, semi-routine, lower supervisory, and long-term unemployed); highest level of education (post-16 qualifications: yes/no); ethnicity (ethnic minority: yes/no); presence of children in the household (yes/no); and history of ≥1 diagnosed mental health condition since the age of 16 (yes/no; assessed in waves up to June 2023 among all participants in England and ∼50% of participants in Wales and Scotland).

Smoking status was assessed by asking participants which of the following best applied to them:

a. ‘I smoke cigarettes (including hand-rolled) every day’
b. ‘I smoke cigarettes (including hand-rolled), but not every day’
c. ‘I do not smoke cigarettes at all, but I do smoke tobacco of some kind (e.g., pipe, cigar or shisha)’
d. ‘I have stopped smoking completely in the last year’
e. ‘I stopped smoking completely more than a year ago’
f. ‘I have never been a smoker (i.e., smoked for a year or more)’

Those who responded a to c were considered current smokers, those who responded d recent (<1y) ex-smokers, e long-term (≥1 year) ex-smokers, and f never-smokers.

### Statistical analysis

Analyses were conducted using R v.4.2.1. The Smoking Toolkit Study uses survey weights to adjust data so that the sample matches the demographic profile of Great Britain on age, social grade, region, housing tenure, ethnicity and working status within sex.^25,26^ The following analyses used weighted data. Data were weighted differently for analyses of mental health conditions, to account for this variable not being assessed among all participants in Wales and Scotland. Missing cases were excluded on a per-analysis basis.

We used logistic regression to estimate monthly time trends in the proportion of adults in Great Britain who report using disposable e-cigarettes, stratified by different sociodemographic characteristics and smoking status. For each of the sociodemographic characteristics and smoking status, we constructed a model with time, the characteristic of interest, and their interaction as predictors, thus allowing for time trends to differ across subgroups. Both time (survey wave) and age were modelled continuously using restricted cubic splines with three knots (placed at earliest, middle and latest survey wave for time and 5, 50, and 95% quantiles for age), to allow the relationship of prevalence with time and age to be flexible and non-linear, while avoiding categorisation. As age was modelled continuously, we displayed estimates for six specific ages (18-, 25-, 35-, 45-, 55-, and 65-year-olds) to illustrate how trends differed across ages. Note that the model used to derive these estimates included data from participants of all ages, not only those who were aged exactly 18, 25, 35, 45, 55, or 65 years. Absolute percentage point changes in prevalence across the whole time-series (calculated as prevalence in August 2023 minus prevalence in January 2021) among each population group are presented, alongside 95% confidence intervals (CI) calculated using bootstrapping. We estimated the total number of adults in Great Britain using disposables in August 2023 – overall, by smoking status, and among 18-24 year-old never smokers specifically – based on the most recent (2021) mid-year population estimates for Great Britain^28^ and smoking prevalence data from the Annual Population Survey^29^ (for current and never smoking; UK) and the Smoking Toolkit Study (for recent and long-term ex-smoking; Great Britain).

After conducting these planned trend analyses, we undertook some additional, unplanned analyses to explore differences by age, smoking status, and mental health in more detail. Specifically, we were interested in the extent to which disposable use had risen among never smokers within different age groups, and whether differences in trends between people with and without a history of mental health conditions were consistent across different smoking statuses. We tested the interaction between time and smoking status stratified by age (18-24, 25-44, ≥45 years) and the interaction between time and history of mental health conditions stratified by smoking status.

We then used descriptive statistics to summarise the prevalence of use of (i) any, (ii) disposable, (iii) refillable, and (iv) pod vaping in 2023, and compare the profiles (i.e., sociodemographic characteristics and smoking status) of current users of these products. We report data for all adult users and separately by smoking status.

### Patient and public involvement

The wider Smoking Toolkit Study is discussed several times a year with a diverse PPI group and the authors regularly attend and present at meetings at which patients and the public are included. Interaction and discussion at these events help shape the broad research priorities and questions. There is also a mechanism for generalised input from the wider public: each month interviewers seek feedback on the questions from all 2,450 respondents, who are representative of the British population. This feedback is limited and usually just relates to understanding of questions and item options. No patients or members of the public were involved in setting the research questions or the outcome measures, nor were they involved in the design and implementation of this specific study. There are no plans to involve patients in dissemination but results will be disseminated to stakeholders in relevant public health bodies and charities.

### Results

A total of 75,268 adults aged ≥16 years were surveyed between January 2021 and August 2023. We excluded 4,979 participants surveyed in England in May, June, or August 2022 (when e-cigarette device type was not assessed). For the trend analyses, we also excluded 2,312 16 and 17 year-olds (because these ages were not included in certain waves), leaving a final sample for analysis of 67,977 adults aged ≥18 years. For the descriptive analyses of vaping prevalence and the profile of vapers in 2023, we included data from the 19,134 adults aged ≥16 years surveyed between January 2023 and August 2023. In total, we analysed data from 69,973 unique participants. Characteristics of the two sub-samples (i.e., those contributing data to the trend analyses and to the 2023 descriptive analyses) are summarised in Table S1.

### Time trends in disposable e-cigarette use

Between January 2021 and August 2023, the prevalence of disposable e-cigarette use among adults aged ≥18 years grew from 0.1% to 4.9% (an absolute rise of 4.85 percentage points [95%CI 4.00-5.15]; Table 1). This equates to approximately 2.6 million adult disposable vapers in Great Britain in August 2023 (53.2 million adults^28^ x 4.9%).

**Table 1.**
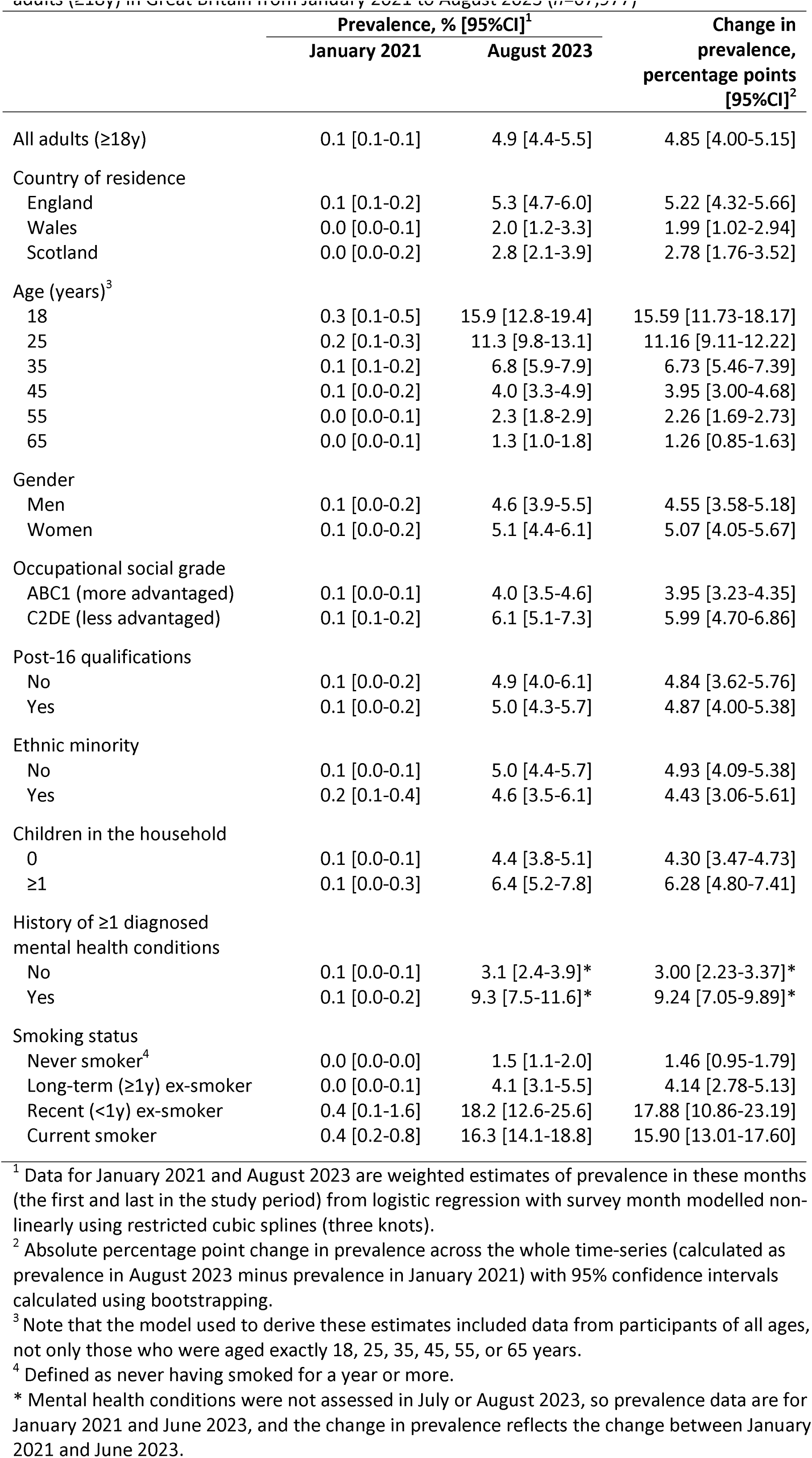
Modelled estimates of the change in prevalence of disposable e-cigarette use among adults (≥18y) in Great Britain from January 2021 to August 2023 (n=67,977)

This rise in prevalence was observed across all population subgroups to varying degrees (Figure 1). In particular, there were substantial differences in the absolute rise in prevalence by age and smoking status (Table 1). Disposable e-cigarette use increased more rapidly, and to much higher levels, among younger than older adults (Figure 1B). For example, the prevalence of disposable use among 18-year-olds increased from 0.3% in January 2021 to 15.9% in August 2023, whereas it increased from 0.07% to 4.0% among 45-year-olds and from 0.05% to 1.3% among 65-year-olds (Table 1). Growth in disposable e-cigarette use was also much more pronounced among current smokers and those who had quit in the past year (Figure 1I), increasing from 0.4% to 16.3% (an estimated 1.1 million current smokers; 53.2 million adults^28^ x 14.0% current smokers^29^ x 16.3%) and from 0.4% to 18.2% (an estimated 242,000 recent ex-smokers; 53.2 million adults^28^ x 2.5% recent ex-smokers x 18.2%), respectively (Table 1). Use of disposables among those who had quit more than a year ago and had never regularly smoked remained relatively rare (<0.3% and <0.2%) throughout 2021, but increased to 4.1% (an estimated 502,000 long-term ex-smokers; 53.2 million adults^28^ x 23.0% long-term ex-smokers x 4.1%) and 1.5% (an estimated 496,000 never smokers; 53.2 million adults^28^ x 62.1% never smokers^29^ x 1.5%) respectively by August 2023 (Table 1). An exploratory analysis showed the rise in disposable use among never smokers was greatest among younger adults: 7.1% of 18-24-year-olds who had never regularly smoked used disposables in August 2023 (an estimated 316,000 people; 5.39 million year-olds^28^ x 82.7% never smokers^29^ x 7.1%), compared with 1.2% of those aged 25-44 and 0.1% of those aged ≥45 (Figure S1).

**Figure 1.**
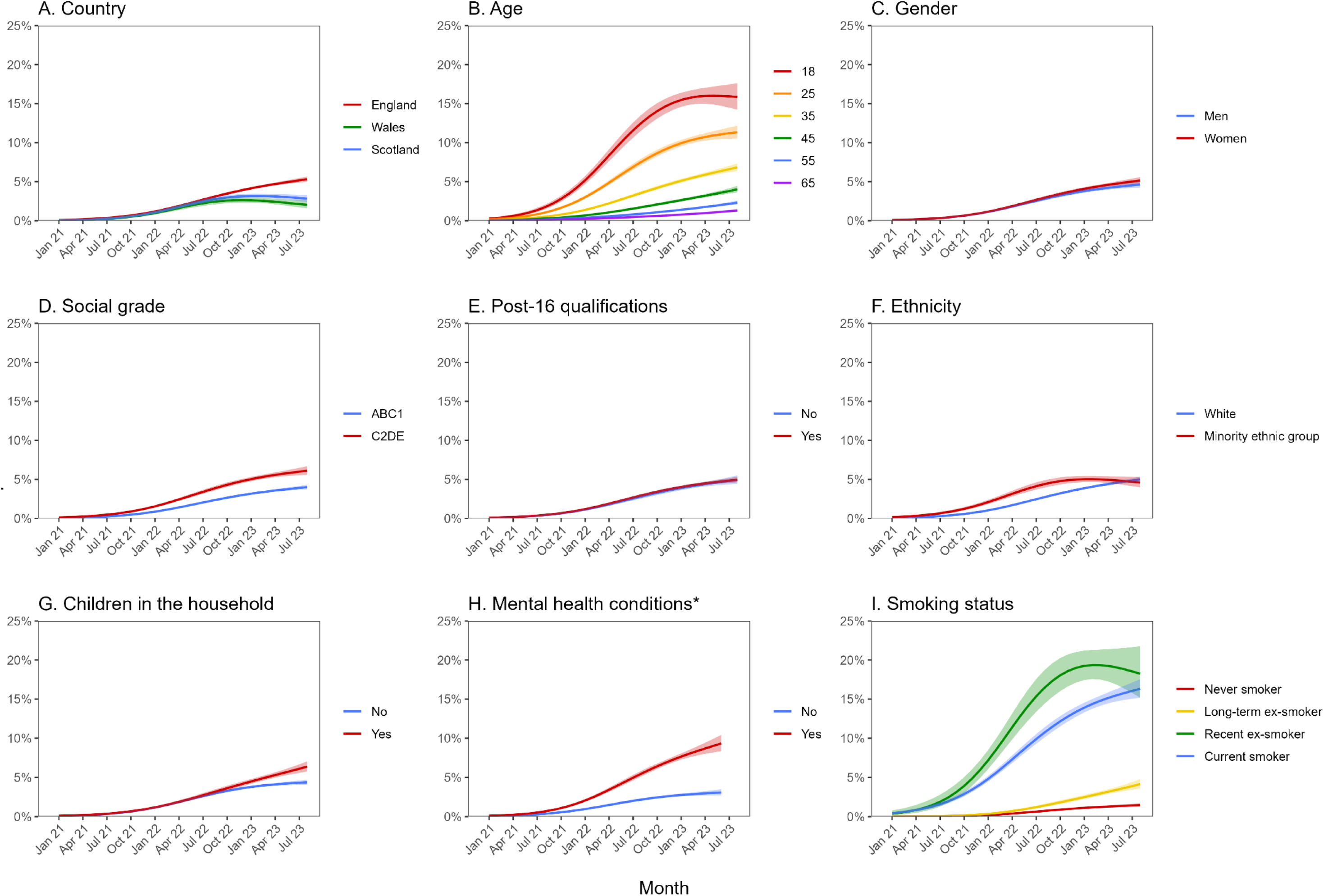
Trends in disposable e-cigarette use among adults (≥18y) in Great Britain, January 2021 to August 2023, by sociodemographic characteristics and smoking status (*n*=67,977). Panels show trends by (A) country of residence, (B) age, (C) gender, (D) occupational social grade, (E) highest level of education, (F) ethnicity, (G) children in the household, (H) history of mental health conditions, and (I) smoking status. Lines represent modelled weighted prevalence by monthly survey wave, modelled non-linearly using restricted cubic splines (three knots). Shaded bands represent standard errors. * Mental health conditions were not assessed in July or August 2023, so trends were modelled up to June 2023.

Trends in disposable e-cigarette use also differed by country of residence and ethnicity. The rise in prevalence was similar across countries up to October 2022, but then levelled off in Wales and Scotland while continuing to rise in England (Figure 1A). As a result, the proportion of adults using disposable e-cigarettes in August 2023 was twice as high in England (5.3%) as in Scotland (2.8%) or Wales (2.0%; Table 1). Disposable use initially grew faster among adults from ethnic minority groups than among white adults, but the rate of increase was similar through 2022 and higher among white adults in 2023 (Figure 1F), such that prevalence was similar by August 2023 (Table 1).

In addition, the rate of increase differed by occupational social grade (Figure 1D), the presence of children in the household (Figure 1G), and history of mental health conditions (Figure 1H). As a result, at the end of the study period, the prevalence of disposable use was significantly higher among less (vs. more) advantaged social grades, those with (vs. without) children, and those with (vs. without) a history of mental health conditions (Table 1). Disposable use was particularly prevalent (9.3%) among adults with a history of mental health conditions in June 2023 (the last wave to assess mental health diagnoses; Table 1). An exploratory analysis stratified by smoking status showed the divergence in trends between those with and without a history of mental health conditions was most pronounced among people who had stopped smoking in the past year: in June 2023, 33.5% of recent ex-smokers with a history of mental health conditions used disposables compared with 10.3% of those without mental health conditions (Figure S2). A similar pattern was observed when age was included as an adjustment (Figure S3), suggesting this was not driven by those who reported a history of mental health conditions being younger, on average, than those who did not (44.9 [SD 17.4] vs. 50.5 [SD 18.6] years).

### Profile of disposable vapers in 2023

Table 2 shows the prevalence of vaping, overall and by device type, among adults (≥16 years) in 2023 (aggregated across January–August). Groups more likely to be using disposables than either refillable or pod e-cigarettes included 16-24-year-olds, ethnic minorities, never smokers, and current smokers.

**Table 2.** Prevalence of vaping, overall and by device type, among adults (≥16y) in Great Britain in 2023 (n=19,134)

|  | Prevalence of vaping, %<sup>1</sup> (95%CI) |  |  |  |
| --- | --- | --- | --- | --- |
|  | Any vaping | Disposable vaping<sup>2</sup> | Refillable vaping | Pod vaping |
| All adults (≥16y) | 11.3 [10.8–11.9] | 4.6 [4.2–5.0] | 5.1 [4.7–5.5] | 1.2 [1.0–1.4] |
| Country of residence |  |  |  |  |
| England | 11.8 [11.2–12.4] | 4.9 [4.5–5.3] | 5.2 [4.8–5.6] | 1.3 [1.1–1.5] |
| Wales | 8.0 [6.6–9.4] | 2.5 [1.7–3.3] | 4.9 [3.8–6.1] | 0.4 [0.1–0.7] |
| Scotland | 8.5 [7.5–9.5] | 3.1 [2.4–3.7] | 4.4 [3.7–5.1] | 0.8 [0.5–1.2] |
| Age (years) |  |  |  |  |
| 16-24 | 23.3 [21.1–25.5] | 14.4 [12.6–16.2] | 6.2 [5.0–7.5] | 1.9 [1.2–2.6] |
| 25-34 | 16.4 [14.8–18.0] | 7.1 [5.9–8.2] | 7.2 [6.1–8.4] | 1.5 [1.0–2.0] |
| 35-44 | 13.4 [11.9–14.9] | 4.7 [3.8–5.7] | 6.7 [5.6–7.9] | 1.4 [0.9–1.9] |
| 45-54 | 9.6 [8.4–10.7] | 2.3 [1.7–2.9] | 5.7 [4.8–6.7] | 1.0 [0.6–1.4] |
| 55-64 | 8.3 [7.2–9.3] | 2.2 [1.6–2.8] | 4.6 [3.8–5.4] | 1.3 [0.9–1.8] |
| ≥65 | 2.6 [2.1–3.1] | 0.4 [0.2–0.6] | 1.7 [1.3–2.1] | 0.5 [0.3–0.7] |
| Gender |  |  |  |  |
| Men | 12.0 [11.1–12.8] | 4.4 [3.9–4.9] | 6.0 [5.4–6.6] | 1.1 [0.9–1.4] |
| Women | 10.4 [9.7–11.2] | 4.7 [4.2–5.3] | 4.2 [3.7–4.7] | 1.2 [0.9–1.4] |
| Other | 26.8 [19.7–34.0] | 10.1 [5.2–14.9] | 10.7 [5.7–15.8] | 3.4 [0.4–6.3] |
| Occupational social grade |  |  |  |  |
| ABC1 (more advantaged) | 9.3 [8.7–9.8] | 3.7 [3.3–4.1] | 4.2 [3.8–4.5] | 1.1 [0.9–1.3] |
| C2DE (less advantaged) | 13.9 [12.9–14.9] | 5.8 [5.1–6.5] | 6.3 [5.6–7.0] | 1.3 [1.0–1.6] |
| Post-16 qualifications |  |  |  |  |
| No | 12.3 [11.3–13.3] | 4.6 [4.0–5.3] | 5.8 [5.1–6.5] | 1.5 [1.1–1.9] |
| Yes | 10.8 [10.2–11.5] | 4.6 [4.2–5.1] | 4.8 [4.4–5.3] | 1.0 [0.8–1.2] |
| Ethnic minority |  |  |  |  |
| No | 11.6 [11.0–12.3] | 4.6 [4.1–5.0] | 5.5 [5.0–5.9] | 1.2 [1.0–1.4] |
| Yes | 9.5 [8.2–10.7] | 5.1 [4.2–6.1] | 3.0 [2.3–3.8] | 1.0 [0.6–1.4] |
| Children in the household |  |  |  |  |
| 0 | 10.6 [10.0–11.2] | 4.2 [3.7–4.6] | 4.8 [4.4–5.2] | 1.2 [1.0–1.4] |
| ≥1 | 13.0 [11.9–14.1] | 5.7 [4.9–6.5] | 5.8 [5.0–6.6] | 1.1 [0.8–1.4] |
| History of ≥1 diagnosed mental health conditions* |  |  |  |  |
| No | 7.7 [7.1–8.4] | 3.1 [2.6–3.5] | 3.4 [3.0–3.8] | 1.0 [0.7–1.2] |
| Yes | 18.5 [17.1–19.9] | 7.9 [6.9–8.9] | 8.1 [7.1–9.1] | 1.6 [1.2–2.0] |
| Smoking status |  |  |  |  |
| Never smoker<sup>3</sup> | 2.7 [2.3–3.1] | 1.5 [1.2–1.8] | 0.9 [0.7–1.1] | 0.2 [0.1–0.3] |
| Long-term (≥1y) ex-smoker | 15.4 [14.1–16.7] | 3.2 [2.5–3.9] | 10.1 [9.0–11.2] | 1.7 [1.3–2.2] |
| Recent (<1y) ex-smoker | 46.0 [40.3–51.7] | 19.0 [14.4–23.7] | 19.7 [15.1–24.2] | 5.5 [2.8–8.2] |
| Current smoker | 31.5 [29.5–33.5] | 15.8 [14.1–17.4] | 11.1 [9.8–12.4] | 3.4 [2.6–4.1] |

<sup>1</sup> Row percentages.
<sup>2</sup> Note these are unmodelled estimates based on aggregated data across January-August 2023, so they differ slightly from the modelled estimates of prevalence in August 2023 presented in **Table 1**.
<sup>3</sup> Defined as never having smoked for a year or more.
\* Mental health conditions were not assessed in July or August 2023, so results are based on aggregated data across January-June 2023.
Corresponding data are provided separately by smoking status in **Tables S2-5**.

Table 3 shows the sociodemographic and smoking characteristics of vapers in 2023, overall and by the main type of device used. Of this group, 42.4% (weighted) were using disposables, 46.8% refillable devices, and 10.9% pod devices.

**Table 3.** Sociodemographic and smoking profile of adult (≥16y) e-cigarette users in Great Britain in 2023 (n=1,842)

|  | All<br>e-cigarette users<br>%<sup>1</sup> (95%CI) | Disposable<br>e-cigarette users<br>%<sup>1</sup> (95%CI) | Refillable<br>e-cigarette users<br>%<sup>1</sup> (95%CI) | Pod<br>e-cigarette users<br>%<sup>1</sup> (95%CI) |
| --- | --- | --- | --- | --- |
| *Unweighted N*<sup>2</sup> | 1,842 | 703 | 878 | 201 |
| Country of residence |  |  |  |  |
| England | 90.0 [88.9–91.1] | 91.6 [89.9–93.1] | 87.8 [85.9–89.5] | 92.4 [89.0–94.8] |
| Wales | 3.5 [2.9–4.2] | 2.6 [1.9–3.6] | 4.7 [3.7–6.0] | 1.6 [0.7–3.5] |
| Scotland | 6.5 [5.7–7.4] | 5.7 [4.6–7.2] | 7.5 [6.3–8.9] | 6.0 [3.9–9.1] |
| Age (years) |  |  |  |  |
| 16-24 | 26.9 [24.6–29.4] | 40.7 [36.6–44.9] | 16.0 [13.2–19.1] | 21.1 [15.0–28.8] |
| 25-34 | 24.1 [21.9–26.5] | 25.5 [22.0–29.3] | 23.5 [20.3–27.1] | 20.6 [15.0–27.6] |
| 35-44 | 18.7 [16.7–20.8] | 16.2 [13.3–19.5] | 20.9 [17.9–24.2] | 18.2 [13.0–25.0] |
| 45-54 | 13.7 [12.0–15.4] | 8.0 [6.1–10.5] | 18.1 [15.5–21.1] | 14.1 [9.7–20.1] |
| 55-64 | 11.3 [9.9–12.9] | 7.4 [5.6–9.8] | 14.0 [11.8–16.5] | 17.2 [12.3–23.4] |
| ≥65 | 5.3 [4.3–6.4] | 2.2 [1.4–3.4] | 7.5 [5.9–9.5] | 8.8 [5.6–13.5] |
| Gender |  |  |  |  |
| Men | 51.4 [48.8–54.0] | 46.4 [42.2–50.6] | 57.2 [53.4–60.9] | 47.2 [39.5–55.0] |
| Women | 46.8 [44.2–49.4] | 51.9 [47.7–56.1] | 41.2 [37.5–45.0] | 50.6 [42.8–58.3] |
| Other | 1.9 [1.4–2.5] | 1.7 [1.0–2.8] | 1.6 [1.0–2.7] | 2.2 [0.9–5.3] |
| Social grade C2DE (less advantaged) | 54.0 [51.5–56.6] | 55.3 [51.2–59.3] | 54.3 [50.6–58.0] | 47.8 [40.1–55.6] |
| Post-16 qualifications | 65.8 [63.3–68.3] | 68.6 [64.6–72.4] | 64.6 [60.8–68.2] | 60.0 [52.0–67.4] |
| Ethnic minority | 11.8 [10.3–13.4] | 15.6 [13.0–18.6] | 8.3 [6.5–10.6] | 11.9 [7.8–17.7] |
| ≥1 children in the household | 33.8 [31.3–36.3] | 36.2 [32.3–40.4] | 33.4 [29.8–37.1] | 27.0 [20.6–34.6] |
| History of ≥1 diagnosed mental health conditions | 53.4 [50.4–56.4] | 55.3 [50.4–60.1] | 53.1 [48.6–57.6] | 43.6 [34.9–52.7] |
| Smoking status |  |  |  |  |
| Never smoker<sup>3</sup> | 13.8 [12.1–15.7] | 19.0 [16.0–22.4] | 10.2 [8.1–12.8] | 9.2 [5.6–14.6] |
| Long-term (≥1y) ex-smoker | 31.1 [28.8–33.6] | 15.8 [12.9–19.2] | 45.2 [41.5–49.0] | 33.6 [26.8–41.2] |
| Recent (<1y) ex-smoker | 9.8 [8.3–11.6] | 9.9 [7.6–12.8] | 9.3 [7.2–11.8] | 11.2 [6.9–17.8] |
| Current smoker | 45.2 [42.7–47.8] | 55.3 [51.1–59.4] | 35.3 [31.7–39.0] | 46.0 [38.4–53.7] |

<sup>1</sup> Column percentages.<sup>2</sup> Note that numbers of disposable, refillable, and pod e-cigarette users do not sum to the number of all e-cigarette users because some participants (*n*=60) did not report the main device type they used.<sup>3</sup> Defined as never having smoked for a year or more.Corresponding data are provided separately by smoking status in **Tables S5-8**.

On average, disposable users were younger than those who used other types of e-cigarettes (Table 3): 40.7% of disposable users were aged 16-24, compared with 16.0% of refillable users and 21.1% of pod users. The proportion aged 25-44 was similar across device types, and the proportion aged ≥45 was lower among disposable users.

There were also notable differences by smoking status (Table 3). The proportion of disposable users who had never been a regular smoker was around double that of refillable and pod users (19.0% vs. 10.2% and 9.2%, respectively). A higher proportion of disposable users were current smokers and a lower proportion were long-term ex-smokers. The proportion of recent ex-smokers was similar across device types.

Disposable users also differed from refillable users (but were similar to pod users) on country of residence, gender and ethnicity (Table 3). A higher proportion of disposable than refillable users lived in England. Higher proportions of disposable than refillable users identified as women and as belonging to ethnic minority groups.

Disposable users did not differ significantly from refillable or pod users on occupational social grade, education, children in the household, or history of mental health conditions (Table 3).

## Discussion

Use of disposable e-cigarettes rose sharply between 2021 and 2023 in Great Britain. In August 2023, one in 20 adults reported using disposables, which equates to approximately 2.6 million current users who would be affected if a disposables ban were to be implemented. While this rise was observed across all population subgroups, it was most pronounced among younger adults, those who currently smoke, and those who had stopped smoking in the past year. Use among those who had never regularly smoked remained relatively rare, but was higher among 18-24 year-olds. Use of disposables has risen to higher levels in England than in Wales or Scotland, and among people from less advantaged social grades, those with children in the household, and those with a history of mental health conditions.

The UK Government is consulting on banning disposable vapes.^12^ This appears to have widespread support from practitioners,^24^ politicians,^22^ and the general public.^30^ Alongside concerns about disposables’ negative environmental impact, a key concern motivating calls for a ban is the rapid rise in vaping among young people – particularly young never smokers – that has been driven by these products.^22,24^ Our data show that adults under the age of 25 and those who have never regularly smoked are more likely to use disposable e-cigarettes than refillable or pod devices, suggesting banning disposables would particularly impact these target groups. We estimate that there are approximately 316,000 people in Great Britain aged between 18 and 24 who have never regularly smoked currently using disposable e-cigarettes. This number likely reflects some people who were diverted from using other nicotine products (e.g., rechargeable e-cigarettes or cigarettes), as well as others who would not have otherwise vaped or smoked.

However, our data also show that a ban would disproportionately affect the ∼1.1 million current smokers who make up around half of the population of disposable users. This is a group who would benefit from harm reduction if they switched completely to e-cigarettes.^21^ It would also affect ∼242,000 people who have recently switched completely from smoking to vaping and ∼502,000 people who quit smoking more than a year ago. In the event of a ban, it would be important to encourage current and ex-smokers who use disposables to switch to other (rechargeable) types of e-cigarettes rather than going back to just smoking tobacco. In particular, it may be recent ex-smokers who are most likely to relapse: if they chose disposables as a preferable quitting method over other types of e-cigarettes, they may be more vulnerable to relapse even if offered other e-cigarette devices. Growth in use among long-term ex-smokers took longer to appear – use remained relatively rare throughout 2021, which contrasted with current and recent ex-smokers – and this pattern is more consistent with people quitting smoking with the help of disposables and accumulating years of abstinence than long-term ex-smokers with many years of abstinence starting to use disposables. Any policies to reduce the appeal of disposables would probably have fewer unintended consequences the sooner they are enacted, and the less time there has been for ever greater numbers of ex-smokers to rely on these specific devices. In addition, we found disposable use to be particularly prevalent among recent ex-smokers with a history of mental health conditions. People with mental health conditions have much higher rates of both smoking and vaping, compared with the general population.^31^ Previous studies have suggested people with severe mental illness may find disposable devices easier to use than other types of e-cigarettes as they do not require charging or replacement of cartridges/pods.^32,33^ If a ban is implemented, this group may require targeted support to help them avoid relapse to smoking.

Whether to ban disposables also requires careful consideration of other potential unintended consequences. While it is likely to reduce the number of young people taking up vaping, banning disposables would not immediately stop those who already vape from doing so. E-cigarettes contain nicotine, which is an addictive substance, and while a ban would likely encourage some to quit vaping, others will be unwilling or unable to stop. Another issue is that a ban will take time to implement^36^ – which is a concern in the context of rapid growth in the prevalence of vaping among young people. In addition, the e-cigarette market is evolving rapidly and e-cigarette manufacturers may be more likely to respond to a ban (than other measures) by adapting their product offering to meet demand. Leading disposable brands have already launched refillable versions with similar branding and flavours, which are sold for ∼£8. This risks a ban only being effective for a short time before new products, which meet legal requirements but are attractive and affordable for young people, are introduced. In addition, a ban may signal to smokers and ex-smokers that stronger measures are warranted for e-cigarettes than cigarettes. Comparative harm perceptions among adults currently smoking are already extremely inaccurate with only 27% believing e-cigarettes are less harmful than ^37^ Vaping is substantially less harmful than smoking,^21^ so it is essential that a ban on disposables does not discourage people who smoke from switching to vaping.

Alternative measures to strengthen the regulation of disposable vapes have been proposed,^18^ which could be implemented more quickly than a ban, adapted more flexibly, and may reduce the risk of causing relapse in ex-smokers who are using the product.^36^ They include: (i) implementing an excise tax on disposables to raise the price to the same level as the cheapest reusable e-cigarettes (making disposables less affordable to children and encouraging adult users to switch to less environmentally damaging products); (ii) prohibiting branding with appeal to children (e.g., bright colours, sweet names, and cartoon characters) across all e-cigarette products and packaging (not just disposables); (iii) prohibiting promotion of e-cigarettes in shops, putting e-cigarettes out of sight and reach of children; and (iv) giving greater powers to Border Force, HMRC, and Trading Standards to control the import, distribution, and sale of disposable e-cigarettes.^18^

Strengths of this study include the large, nationally representative sample and monthly data collection, which provides granular and up-to-date estimates of prevalence. There were also several limitations. Participants aged 16 and 17 were not surveyed in 2021, so we were unable to include these ages in the trend analyses. The measure of disposable e-cigarette use did not distinguish modern disposables from older ‘cigalikes’. Moreover, it asked about which type of e-cigarette vapers mainly use, so vapers who used disposables as a secondary product were not captured. This may be a particular issue for assessing disposable use as people who mainly use other e-cigarette devices may buy them as temporary replacements if they forget to take their device out with them or if it runs out of battery while away from home. As a result, the estimated prevalence of disposable vaping actually represents a lower bound for the true prevalence. In addition, the measure of smoking status defines being a never smoker as not having ‘smoked for a year or more’ so this group includes those who have regularly smoked, but for less than a year. This means the estimated prevalence of disposable vaping among never smokers likely overestimates the true prevalence among people who have never regularly smoked. Nonetheless, these data provide the most comprehensive and up-to-date information on disposable e-cigarette use in Great Britain and provide useful information to inform decision-making about how to regulate these products. They offer insight into the number and profile of people who would currently be affected, although we note that a ban would also affect people who may have started using the products in the future if they were available – which, if current trends in disposable vaping continue, will add up to a substantial number. It will be important to continue to monitor disposable use as the market continues to evolve rapidly.

In conclusion, a ban on disposable e-cigarettes would currently affect approximately 2.6 million adults in Great Britain. The impact of a ban would be greatest for young people, including the 316,000 18-24 year-olds who have never regularly smoked tobacco, which may make it an appealing option to policymakers and the general public. However, banning disposables would also affect around 1.1 million people who currently smoke and a further 744,000 who previously smoked. In addition, it would have a disproportionate impact on disadvantaged groups that have higher rates of smoking and typically find it harder to quit. As such, it has the potential to slow progress in driving down smoking prevalence, reducing smoking-related harm (through switching from smoking to vaping) and could worsen health inequalities. These trade-offs, as well as other potential unintended consequences, require careful consideration when weighing the benefits of an outright ban on disposables against other regulatory policies.

## Declarations

### Ethics approval

Ethical approval for the STS was granted originally by the UCL Ethics Committee (ID 0498/001). The data are not collected by UCL and are anonymized when received by UCL.

## Data Availability

Data are available on Open Science Framework (https://osf.io/uwfd3/).

