## Supplementary material for "Who would be affected by a ban on disposable vapes? A population study in Great Britain"

##### Table S1. Weighted sample characteristics

|  | **Trend analyses:**  **adults aged ≥18 years**  **(*n*^1^=67,977)** | **Descriptive analyses (2023): adults aged ≥16 years**  **(*n*^1^=19,134)** |
| --- | --- | --- |
| Country of residence |  |  |
| England | 85.2% | 86.4% |
| Wales | 5.3% | 4.9% |
| Scotland | 9.5% | 8.6% |
| Age (years) |  |  |
| Mean (SD) | 48.8 (18.5) | 48.0 (18.6) |
| 16-24 | 11.5% | 13.1% |
| 25-34 | 17.1% | 16.6% |
| 35-44 | 15.8% | 15.8% |
| 45-54 | 16.7% | 16.1% |
| 55-64 | 15.4% | 15.5% |
| ≥65 | 23.4% | 22.8% |
| Gender |  |  |
| Men | 48.6% | 48.6% |
| Women | 50.7% | 50.6% |
| Other | 0.7% | 0.8% |
| Missing, *n***^1^** | 145 | 104 |
| Occupational social grade |  |  |
| ABC1 (more advantaged) | 55.9% | 66.0% |
| C2DE (less advantaged) | 44.1% | 44.0% |
| Post-16 qualifications |  |  |
| No | 32.7% | 31.4% |
| Yes | 67.3% | 68.6% |
| Missing, *n***^1^** | 755 | 271 |
| Ethnic minority |  |  |
| No | 86.9% | 85.9% |
| Yes | 13.1% | 14.1% |
| Missing, *n***^1^** | 531 | 209 |
| Children in the household |  |  |
| 0 | 71.6% | 70.7% |
| ≥1 | 28.4% | 29.3% |
| History of ≥1 diagnosed mental health conditions^2^ |  |  |
| No | 68.6% | 67.5% |
| Yes | 31.4% | 32.5% |
| Missing, *n***^1^** | 9,152 | 6,778 |
| Smoking status |  |  |
| Never smoker | 57.7% | 58.3% |
| Long-term (≥1y) ex-smoker | 23.2% | 23.0% |
| Recent (<1y) ex-smoker | 2.5% | 2.4% |
| Current smoker | 16.6% | 16.3% |
| Missing, *n***^1^** | 519 | 114 |

^1^ Unweighted sample size.

^2^ Mental health conditions were not assessed in July or August 2023; the number of missing cases does not include participants surveyed in these waves. This variable was only assessed in ~50% of participants surveyed in Wales and Scotland. Survey weights are applied to account for this in our analyses.

Note: Data are shown as weighted column percentages, unless otherwise specified. There were some missing data (unweighted *n*s indicated in the table); valid percentages are shown for ease of interpretation.

##### Table S2. Prevalence of vaping, overall and by device type, among adults (≥16y) in Great Britain in 2023: never smokers (*n*=11,224)

|  | **Prevalence, %^1^ (95%CI)** | | | |
| --- | --- | --- | --- | --- |
|  | **Any**  **vaping** | **Disposable**  **vaping** | **Refillable**  **vaping** | **Pod**  **vaping** |
| All adults (≥16y) | 2.7 [2.3–3.1] | 1.5 [1.2–1.8] | 0.9 [0.7–1.1] | 0.2 [0.1–0.3] |
| Country of residence |  |  |  |  |
| England | 2.9 [2.4–3.3] | 1.6 [1.3–1.9] | 0.9 [0.7–1.2] | 0.2 [0.1–0.3] |
| Wales | 1.7 [0.8–2.7] | 1.0 [0.3–1.7] | 0.7 [0.1–1.2] | 0.1 [0.0–0.3] |
| Scotland | 1.6 [1.0–2.3] | 0.8 [0.3–1.3] | 0.6 [0.2–1.1] | 0.0 [0.0–0.1] |
| Age (years) |  |  |  |  |
| 16-24 | 9.6 [7.9–11.4] | 6.7 [5.3–8.2] | 1.9 [1.1–2.6] | 0.5 [0.1–0.9] |
| 25-34 | 3.0 [2.0–4.0] | 1.3 [0.7–1.9] | 1.3 [0.6–2.0] | 0.3 [0.0–0.7] |
| 35-44 | 2.5 [1.6–3.4] | 1.2 [0.5–1.8] | 1.1 [0.5–1.6] | 0.2 [0.0–0.4] |
| 45-54 | 0.9 [0.3–1.5] | 0.0 [0.0–0.1] | 0.9 [0.3–1.4] | 0.0 [0.0–0.1] |
| 55-64 | 0.8 [0.3–1.2] | 0.3 [0.0–0.6] | 0.5 [0.1–0.8] | 0.0 [0.0–0.0] |
| ≥65 | 0.2 [0.0–0.3] | 0.0 [0.0–0.1] | 0.1 [0.0–0.2] | 0.0 [0.0–0.1] |
| Gender |  |  |  |  |
| Men | 3.0 [2.4–3.5] | 1.4 [1.0–1.8] | 1.2 [0.8–1.5] | 0.2 [0.1–0.4] |
| Women | 2.4 [1.9–2.9] | 1.6 [1.2–2.0] | 0.6 [0.4–0.9] | 0.1 [0.0–0.2] |
| Occupational social grade |  |  |  |  |
| ABC1 (more advantaged) | 2.2 [1.8–2.5] | 1.2 [1.0–1.5] | 0.7 [0.5–0.9] | 0.2 [0.1–0.3] |
| C2DE (less advantaged) | 3.5 [2.7–4.3] | 1.9 [1.4–2.5] | 1.3 [0.8–1.7] | 0.2 [0.0–0.4] |
| Post-16 qualifications |  |  |  |  |
| No | 2.9 [2.1–3.6] | 1.3 [0.8–1.8] | 1.2 [0.7–1.7] | 0.2 [0.0–0.4] |
| Yes | 2.7 [2.2–3.1] | 1.6 [1.3–1.9] | 0.8 [0.6–1.1] | 0.2 [0.1–0.3] |
| Ethnic minority |  |  |  |  |
| No | 2.5 [2.1–2.9] | 1.3 [1.0–1.6] | 1.0 [0.7–1.2] | 0.2 [0.1–0.3] |
| Yes | 3.6 [2.6–4.6] | 2.7 [1.8–3.6] | 0.5 [0.1–0.9] | 0.1 [0.0–0.3] |
| Children in the household |  |  |  |  |
| 0 | 2.5 [2.0–2.9] | 1.3 [1.0–1.7] | 0.9 [0.6–1.1] | 0.2 [0.1–0.3] |
| ≥1 | 3.2 [2.5–3.9] | 1.9 [1.4–2.5] | 1.0 [0.6–1.4] | 0.2 [0.1–0.4] |
| History of ≥1 diagnosed mental health conditions |  |  |  |  |
| No | 2.0 [1.6–2.5] | 1.1 [0.8–1.4] | 0.6 [0.3–0.9] | 0.2 [0.0–0.3] |
| Yes | 4.7 [3.6–5.7] | 2.9 [2.1–3.8] | 1.6 [1.0–2.2] | 0.2 [0.0–0.4] |

^1^ Row percentages.

##### Table S3. Prevalence of vaping, overall and by device type, among adults (≥16y) in Great Britain in 2023: long-term (≥1 year) ex-smokers (*n*=4,629)

|  | **Prevalence, %^1^ (95%CI)** | | | |
| --- | --- | --- | --- | --- |
|  | **Any**  **vaping** | **Disposable**  **vaping** | **Refillable**  **vaping** | **Pod**  **vaping** |
| All adults (≥16y) | 15.4 [14.1–16.7] | 3.2 [2.5–3.9] | 10.1 [9.0–11.2] | 1.7 [1.3–2.2] |
| Country of residence |  |  |  |  |
| England | 15.9 [14.4–17.3] | 3.4 [2.6–4.2] | 10.2 [9.0–11.4] | 1.9 [1.4–2.5] |
| Wales | 13.0 [9.6–16.5] | 2.2 [0.6–3.8] | 9.9 [6.9–13.0] | 0.3 [0.0–0.7] |
| Scotland | 12.5 [10.2–14.8] | 2.0 [1.0–3.1] | 9.2 [7.2–11.2] | 0.9 [0.2–1.6] |
| Age (years) |  |  |  |  |
| 16-24 | 50.2 [38.6–61.9] | 20.7 [11.1–30.4] | 25.3 [14.7–35.9] | 3.5 [0.0–7.5] |
| 25-34 | 32.3 [27.0–37.6] | 9.8 [6.3–13.3] | 20.1 [15.5–24.7] | 2.1 [0.6–3.6] |
| 35-44 | 23.6 [19.6–27.7] | 4.7 [2.7–6.6] | 15.4 [11.9–18.8] | 2.6 [1.0–4.2] |
| 45-54 | 18.2 [15.1–21.3] | 2.6 [1.2–4.0] | 13.1 [10.4–15.8] | 1.9 [0.8–3.1] |
| 55-64 | 12.1 [9.8–14.5] | 1.2 [0.2–2.2] | 8.3 [6.4–10.2] | 2.5 [1.3–3.6] |
| ≥65 | 3.7 [2.7–4.6] | 0.3 [0.0–0.5] | 2.7 [1.9–3.6] | 0.6 [0.2–1.0] |
| Gender |  |  |  |  |
| Men | 16.0 [14.2–17.8] | 3.0 [2.1–3.9] | 11.6 [10.1–13.2] | 1.2 [0.7–1.7] |
| Women | 14.5 [12.7–16.4] | 3.4 [2.4–4.4] | 8.5 [7.0–10.0] | 2.2 [1.5–3.0] |
| Occupational social grade |  |  |  |  |
| ABC1 (more advantaged) | 13.0 [11.6–14.3] | 2.3 [1.7–2.9] | 8.7 [7.5–9.8] | 1.7 [1.2–2.2] |
| C2DE (less advantaged) | 18.3 [16.0–20.6] | 4.3 [3.0–5.6] | 11.9 [10.0–13.8] | 1.8 [1.1–2.6] |
| Post-16 qualifications |  |  |  |  |
| No | 14.5 [12.4–16.5] | 2.6 [1.6–3.6] | 9.5 [7.7–11.2] | 1.9 [1.1–2.7] |
| Yes | 16.0 [14.4–17.7] | 3.5 [2.6–4.4] | 10.6 [9.2–12.0] | 1.6 [1.1–2.2] |
| Ethnic minority |  |  |  |  |
| No | 15.5 [14.2–16.9] | 3.2 [2.5–3.9] | 10.3 [9.2–11.4] | 1.7 [1.3–2.2] |
| Yes | 14.7 [10.1–19.4] | 4.0 [1.6–6.4] | 8.3 [4.7–12.0] | 2.4 [0.2–4.5] |
| Children in the household |  |  |  |  |
| 0 | 13.2 [11.9–14.6] | 2.6 [1.9–3.3] | 8.7 [7.6–9.8] | 1.6 [1.1–2.1] |
| ≥1 | 21.8 [18.8–24.8] | 5.0 [3.3–6.6] | 14.2 [11.7–16.8] | 2.1 [1.1–3.2] |
| History of ≥1 diagnosed mental health conditions |  |  |  |  |
| No | 10.5 [9.0–12.0] | 1.5 [0.9–2.1] | 7.5 [6.2–8.7] | 1.4 [0.8–2.0] |
| Yes | 22.4 [19.4–25.5] | 5.0 [3.3–6.8] | 14.1 [11.5–16.7] | 2.4 [1.4–3.5] |

^1^ Row percentages.

##### Table S4. Prevalence of vaping, overall and by device type, among adults (≥16y) in Great Britain in 2023: recent (<1 year) ex-smokers (*n*=388)

|  | **Prevalence, %^1^ (95%CI)** | | | |
| --- | --- | --- | --- | --- |
|  | **Any**  **vaping** | **Disposable**  **vaping** | **Refillable**  **vaping** | **Pod**  **vaping** |
| All adults (≥16y) | 46 [40.3–51.7] | 19.0 [14.4–23.7] | 19.7 [15.1–24.2] | 5.5 [2.8–8.2] |
| Country of residence |  |  |  |  |
| England | 47.4 [41.2–53.5] | 19.5 [14.5–24.5] | 20.2 [15.3–25.1] | 5.7 [2.8–8.6] |
| Wales | 32.4 [8.9–55.9] | 16.4 [0.0–33.6] | 16 [0.0–34.5] | 0.0 [0.0–0.0] |
| Scotland | 32.0 [18.6–45.4] | 13.6 [3.4–23.9] | 13.6 [4.1–23.1] | 4.7 [0.0–11.5] |
| Age (years) |  |  |  |  |
| 16-24 | 64.5 [50.0–79.1] | 36.9 [21.8–52.1] | 15.6 [5.1–26.1] | 12.0 [1.3–22.8] |
| 25-34 | 45.3 [35.3–55.4] | 19.5 [11.6–27.3] | 16.4 [9.2–23.5] | 5.3 [0.9–9.7] |
| 35-44 | 46.0 [32.4–59.6] | 15.7 [6.5–25.0] | 25.5 [12.9–38.0] | 3.0 [0.0–6.6] |
| 45-54 | 37.7 [22.5–52.8] | 9.5 [0.0–19.3] | 22.3 [9.4–35.3] | 5.8 [0.0–12.6] |
| 55-64 | 39.8 [24.4–55.2] | 14.7 [2.9–26.5] | 22.5 [8.8–36.1] | 1.3 [0.0–4.0] |
| ≥65 | 26.3 [10.1–42.5] | 3.1 [0.0–6.9] | 20 [4.5–35.6] | 3.1 [0.0–9.4] |
| Gender |  |  |  |  |
| Men | 36.7 [28.9–44.6] | 13.6 [8.2–19.0] | 16.0 [10.1–21.9] | 6.0 [2.1–9.8] |
| Women | 54.7 [46.8–62.6] | 24.2 [16.9–31.6] | 23.2 [16.2–30.1] | 4.8 [1.0–8.6] |
| Occupational social grade |  |  |  |  |
| ABC1 (more advantaged) | 44.5 [37.6–51.3] | 17.5 [12.2–22.7] | 18.9 [13.4–24.4] | 6.5 [3.2–9.9] |
| C2DE (less advantaged) | 47.5 [38.5–56.4] | 20.4 [13.0–27.9] | 20.3 [13.2–27.5] | 4.6 [0.4–8.8] |
| Post-16 qualifications |  |  |  |  |
| No | 46.4 [36.0–56.8] | 19.0 [10.8–27.1] | 20.8 [12.2–29.4] | 4.7 [0.0–9.5] |
| Yes | 45.6 [38.6–52.5] | 19.3 [13.5–25.2] | 18.6 [13.2–23.9] | 6.1 [2.7–9.5] |
| Ethnic minority |  |  |  |  |
| No | 48.2 [41.9–54.5] | 19.0 [13.9–24.1] | 21.8 [16.7–27.0] | 5.6 [2.5–8.6] |
| Yes | 32.5 [19.3–45.8] | 18.8 [7.8–29.9] | 8.2 [0.0–16.3] | 4.5 [0.0–9.6] |
| Children in the household |  |  |  |  |
| 0 | 43.9 [37.1–50.7] | 18.4 [12.9–23.9] | 17.3 [12.2–22.4] | 6.1 [2.6–9.5] |
| ≥1 | 51.3 [40.8–61.8] | 20.5 [11.9–29.2] | 25.4 [16.1–34.7] | 4.2 [0.2–8.1] |
| History of ≥1 diagnosed mental health conditions |  |  |  |  |
| No | 31.4 [23.1–39.7] | 14.4 [8.0–20.7] | 10.0 [5.1–14.9] | 6.3 [1.8–10.8] |
| Yes | 62.5 [53.5–71.4] | 26.7 [18.2–35.3] | 27.4 [18.8–36.0] | 4.5 [0.2–8.8] |

^1^ Row percentages.

##### Table S5. Prevalence of vaping, overall and by device type, among adults (≥16y) in Great Britain in 2023: current smokers (*n*=2,779)

|  | **Prevalence, %^1^ (95%CI)** | | | |
| --- | --- | --- | --- | --- |
|  | **Any**  **vaping** | **Disposable**  **vaping** | **Refillable**  **vaping** | **Pod**  **vaping** |
| All adults (≥16y) | 31.5 [29.5–33.5] | 15.8 [14.1–17.4] | 11.1 [9.8–12.4] | 3.4 [2.6–4.1] |
| Country of residence |  |  |  |  |
| England | 32.0 [29.8–34.2] | 16.2 [14.4–18.0] | 11.0 [9.5–12.4] | 3.4 [2.6–4.2] |
| Wales | 25.3 [19.0–31.5] | 9.1 [5.4–12.8] | 13.9 [8.7–19.2] | 2.0 [0.0–4.1] |
| Scotland | 28.5 [23.7–33.2] | 13.8 [10.0–17.5] | 11.0 [7.8–14.1] | 3.6 [1.7–5.5] |
| Age (years) |  |  |  |  |
| 16-24 | 55.9 [50.3–61.6] | 35.1 [29.7–40.4] | 14.7 [10.8–18.5] | 4.5 [2.3–6.8] |
| 25-34 | 35.4 [30.8–40.0] | 18.3 [14.5–22.0] | 12.3 [9.1–15.5] | 3.2 [1.7–4.8] |
| 35-44 | 31.7 [26.7–36.6] | 14.9 [11.0–18.9] | 11.9 [8.6–15.2] | 3.4 [1.6–5.1] |
| 45-54 | 22.9 [18.8–27.1] | 8.9 [6.1–11.8] | 9.3 [6.5–12.2] | 2.6 [1.1–4.0] |
| 55-64 | 24.1 [19.8–28.4] | 9.1 [6.1–12.0] | 10.7 [7.6–13.7] | 3.9 [1.9–6.0] |
| ≥65 | 11.5 [8.0–15.0] | 3.4 [1.4–5.4] | 5.9 [3.3–8.5] | 2.2 [0.7–3.8] |
| Gender |  |  |  |  |
| Men | 31.5 [28.8–34.3] | 14.6 [12.4–16.8] | 12.4 [10.5–14.3] | 3.2 [2.2–4.2] |
| Women | 30.9 [27.9–33.8] | 17.0 [14.5–19.5] | 9.4 [7.5–11.2] | 3.5 [2.4–4.6] |
| Occupational social grade |  |  |  |  |
| ABC1 (more advantaged) | 32.7 [30.1–35.3] | 16.3 [14.2–18.4] | 11.3 [9.6–13.1] | 3.8 [2.7–4.9] |
| C2DE (less advantaged) | 30.6 [27.7–33.5] | 15.3 [13.0–17.7] | 10.9 [9.0–12.8] | 3.0 [2.0–4.0] |
| Post-16 qualifications |  |  |  |  |
| No | 29.7 [26.4–32.9] | 14.3 [11.8–16.9] | 10.9 [8.6–13.1] | 4.0 [2.6–5.4] |
| Yes | 32.8 [30.3–35.4] | 17.0 [14.8–19.1] | 11.5 [9.8–13.2] | 3.0 [2.2–3.9] |
| Ethnic minority |  |  |  |  |
| No | 31.2 [29.0–33.3] | 15.6 [13.8–17.3] | 10.9 [9.5–12.3] | 3.3 [2.5–4.1] |
| Yes | 35.0 [29.4–40.6] | 17.5 [13.2–21.9] | 12.6 [8.6–16.6] | 4.2 [1.8–6.7] |
| Children in the household |  |  |  |  |
| 0 | 30.4 [28.1–32.7] | 14.4 [12.6–16.2] | 11.1 [9.6–12.7] | 3.6 [2.7–4.5] |
| ≥1 | 34.2 [30.3–38.2] | 19.0 [15.6–22.4] | 11.0 [8.4–13.6] | 2.7 [1.4–4.0] |
| History of ≥1 diagnosed mental health conditions |  |  |  |  |
| No | 28.0 [24.9–31.1] | 14.1 [11.6–16.5] | 9.7 [7.7–11.6] | 3.5 [2.2–4.7] |
| Yes | 36.3 [32.7–39.9] | 18.3 [15.4–21.3] | 12.5 [10.1–15.0] | 3.2 [2.0–4.3] |

^1^ Row percentages.

##### Table S5. Sociodemographic and smoking profile of adult (≥16y) e-cigarette users in Great Britain in 2023: never smokers (*n*=245)

|  | **All**  **e-cigarette users**  **%^1^ (95%CI)** | **Disposable**  **e-cigarette users**  **%^1^ (95%CI)** | **Refillable**  **e-cigarette users**  **%^1^ (95%CI)** | **Pod**  **e-cigarette users**  **%^1^ (95%CI)** |
| --- | --- | --- | --- | --- |
| *Unweighted N* | *245* | *134* | *84* | *18* |
| Country of residence |  |  |  |  |
| England | 91.3 [88.1–93.8] | 92.0 [87.4–95.0] | 89.8 [83.1–94.0] | 95.3 [79.7-99.0] |
| Wales | 3.3 [1.9–5.6] | 3.3 [1.6–6.8] | 3.8 [1.6–8.7] | 2.4 [0.3-18.8] |
| Scotland | 5.4 [3.6–8.0] | 4.7 [2.6–8.4] | 6.4 [3.2–12.1] | 2.3 [0.3-18.3] |
| Age (years) |  |  |  |  |
| 16-24 | 56.2 [49.1–63.1] | 69.9 [60.6–77.8] | 32.5 [22.1–45.0] | 43.7 [20.0-70.6] |
| 25-34 | 18.4 [13.5–24.6] | 14.3 [9.0–21.8] | 23.8 [14.3–36.9] | 30.5 [11.3-60.3] |
| 35-44 | 14.3 [10.0–19.9] | 12.1 [7.0–20.3] | 18.6 [11.1–29.5] | 17.8 [5.6-44.1] |
| 45-54 | 5.4 [2.9–9.9] | 0.2 [0.0–1.7] | 15.0 [7.8–27.0] | 3.8 [0.4-26.9] |
| 55-64 | 4.3 [2.4–7.6] | 2.9 [1.0–7.9] | 7.9 [3.8–15.9] | * |
| ≥65 | 1.4 [0.6–3.1] | 0.6 [0.1–2.4] | 2.1 [0.6–6.7] | 4.2 [0.5-29.2] |
| Gender |  |  |  |  |
| Men | 51.1 [44.0–58.2] | 44.0 [34.8–53.5] | 60.2 [47.6–71.5] | 63.0 [35.7-84.0] |
| Women | 46.9 [39.8-54.0] | 54.3 [44.7-63.5] | 37.8 [26.7-50.4] | 31.9 [13.0-59.5] |
| Other | 2.0 [0.9-4.5] | 1.8 [0.6-5.5] | 2.0 [0.5-8.0] | 5.1 [0.6-33.8] |
| Social grade C2DE (less advantaged) | 51.5 [44.4–58.4] | 50.7 [41.3–60.1] | 55.0 [43.0–66.6] | 40.2 [17.2-68.6] |
| Post-16 qualifications | 71.3 [64.3–77.4] | 76.5 [67.5–83.6] | 65.3 [52.5–76.2] | 70.7 [40.4-89.6] |
| Ethnic minority | 23.9 [18.5–30.4] | 31.7 [23.7–41.0] | 9.9 [4.3–20.9] | 14.3 [3.7-42.3] |
| ≥1 children in the household | 36.6 [30.2–43.5] | 39.2 [30.5–48.6] | 33.9 [23.4–46.2] | 35.3 [15.2-62.4] |
| History of ≥1 diagnosed mental health conditions | 45.7 [37.8–53.9] | 49.5 [38.9–60.1] | 48.9 [34.5–63.4] | 26.7 [6.3-66.3] |

^1^ Column percentages.

* Empty cell: no participants within this group of e-cigarette users fell into this subgroup.

##### Table S6. Sociodemographic and smoking profile of adult (≥16y) e-cigarette users in Great Britain in 2023: long-term (≥1 year) ex-smokers (*n*=610)

|  | **All**  **e-cigarette users**  **%^1^ (95%CI)** | **Disposable**  **e-cigarette users**  **%^1^ (95%CI)** | **Refillable**  **e-cigarette users**  **%^1^ (95%CI)** | **Pod**  **e-cigarette users**  **%^1^ (95%CI)** |
| --- | --- | --- | --- | --- |
| *Unweighted N* | *610* | *108* | *417* | *70* |
| Country of residence |  |  |  |  |
| England | 87.5 [85.3–89.5] | 90.2 [84.8–93.8] | 85.9 [82.9–88.4] | 94.1 [88.2–97.1] |
| Wales | 4.7 [3.5–6.3] | 3.8 [1.8–8.0] | 5.5 [3.9–7.6] | 0.9 [0.2–3.8] |
| Scotland | 7.8 [6.3–9.5] | 6.0 [3.4–10.2] | 8.7 [6.8–11.0] | 5.0 [2.3–10.8] |
| Age (years) |  |  |  |  |
| 16-24 | 9.8 [7.1–13.4] | 19.5 [11.9–30.2] | 7.5 [4.7–11.9] | 6.1 [2.0–17.5] |
| 25-34 | 23.4 [19.6–27.7] | 34.2 [24.6–45.3] | 22.2 [17.6–27.5] | 13.7 [6.8–25.5] |
| 35-44 | 22.9 [19.2–27.1] | 21.7 [14.4–31.4] | 22.6 [18.1–27.9] | 22.4 [12.6–36.6] |
| 45-54 | 21.5 [18.0–25.4] | 14.9 [8.7–24.3] | 23.5 [19.2–28.4] | 20.1 [11.1–33.5] |
| 55-64 | 14.1 [11.6–17.2] | 6.7 [2.8–15.1] | 14.8 [11.7–18.5] | 25.4 [16.1–37.5] |
| ≥65 | 8.3 [6.3–10.7] | 2.9 [1.2–7.2] | 9.4 [6.8–12.7] | 12.4 [6.5–22.3] |
| Gender |  |  |  |  |
| Men | 53.7 [49.0–58.2] | 48.4 [37.7–59.3] | 59.3 [53.6–64.7] | 36.4 [24.9–49.8] |
| Women | 45.2 [40.6-49.8] | 50.1 [39.4-60.9] | 40.3 [34.9-46.0] | 61.0 [47.6-72.8] |
| Other | 1.2 [0.6-2.4] | 1.4 [0.3-5.7] | 0.5 [0.1-1.8] | 2.6 [0.6-10.3] |
| Social grade C2DE (less advantaged) | 53.7 [49.2–58.1] | 60.6 [50.4–70.0] | 53.0 [47.4–58.4] | 47.2 [34.2–60.6] |
| Post-16 qualifications | 65.0 [60.5–69.3] | 69.3 [58.3–78.4] | 65.2 [59.6–70.4] | 58.6 [44.9–71.1] |
| Ethnic minority | 5.7 [4.1–7.9] | 7.4 [4.0–13.5] | 4.9 [3.1–7.6] | 8.1 [3.2–18.8] |
| ≥1 children in the household | 36.0 [31.6–40.6] | 39.5 [29.5–50.5] | 35.8 [30.5–41.4] | 31.1 [19.8–45.2] |
| History of ≥1 diagnosed mental health conditions | 52.6 [47.1–58.1] | 63.8 [50.2–75.5] | 49.7 [43–56.4] | 47.1 [32.6–62.2] |

^1^ Column percentages.

##### Table S7. Sociodemographic and smoking profile of adult (≥16y) e-cigarette users in Great Britain in 2023: recent (<1 year) ex-smokers (*n*=167)

|  | **All**  **e-cigarette users**  **%^1^ (95%CI)** | **Disposable**  **e-cigarette users**  **%^1^ (95%CI)** | **Refillable**  **e-cigarette users**  **%^1^ (95%CI)** | **Pod**  **e-cigarette users**  **%^1^ (95%CI)** |
| --- | --- | --- | --- | --- |
| *Unweighted N* | *167* | *69* | *72* | *20* |
| Country of residence |  |  |  |  |
| England | 93.9 [90.5–96.1] | 93.4 [87.2–96.7] | 93.6 [88.0–96.7] | 94.1 [74.5-98.9] |
| Wales | 1.4 [0.6–3.0] | 1.7 [0.6–4.8] | 1.6 [0.5–5.1] | * |
| Scotland | 4.8 [2.8–8.0] | 4.9 [2.1–11.0] | 4.8 [2.2–10.1] | 5.9 [1.1-25.5] |
| Age (years) |  |  |  |  |
| 16-24 | 25.9 [18.5–35.0] | 35.8 [23.1–50.9] | 14.6 [7.3–27.2] | 40.3 [16.9-69.1] |
| 25-34 | 31.0 [23.8–39.3] | 32.2 [21.2–45.7] | 26.3 [16.7–38.7] | 30.3 [12.1-57.8] |
| 35-44 | 19.5 [13.4–27.4] | 16.1 [8.7–28.0] | 25.3 [14.9–39.5] | 10.6 [2.7-33.3] |
| 45-54 | 8.9 [5.4–14.3] | 5.4 [1.8–15.1] | 12.3 [6.4–22.4] | 11.5 [3.1-34.8] |
| 55-64 | 10.2 [6.2–16.5] | 9.2 [3.8–20.4] | 13.6 [6.8–25.2] | 2.8 [0.3-21.3] |
| ≥65 | 4.4 [2.1–8.9] | 1.3 [0.4–4.2] | 7.9 [3.3–18.0] | 4.4 [0.5-29.9] |
| Gender |  |  |  |  |
| Men | 38.3 [30.4–46.8] | 34.3 [23.0–47.8] | 39.1 [27.3–52.2] | 51.9 [26.3-76.6] |
| Women | 60.3 [51.8-68.3] | 64.6 [51.1-76.1] | 59.8 [46.7-71.7] | 44.1 [20.3-71.0] |
| Other | 1.4 [0.4-4.4] | 1.1 [0.2-8.1] | 1.1 [0.1-7.8] | 3.9 [0.4-27.4] |
| Social grade C2DE (less advantaged) | 54.2 [45.7–62.4] | 56.4 [43.0–68.9] | 54.4 [41.5–66.7] | 43.8 [19.4-71.6] |
| Post-16 qualifications | 68.1 [59.3–75.8] | 68.9 [55.0–80.1] | 66.0 [52.1–77.5] | 73.9 [43.4-91.2] |
| Ethnic minority | 10.1 [6.3–15.9] | 14.1 [7.4–25.3] | 5.9 [2.1–15.5] | 11.7 [3.2-35.0] |
| ≥1 children in the household | 32.5 [25.0–41.0] | 31.5 [20.3–45.3] | 37.7 [25.9–51.1] | 22.1 [7.7-49.1] |
| History of ≥1 diagnosed mental health conditions | 67.4 [58.0–75.5] | 65.9 [50.9–78.3] | 74.0 [60.0–84.4] | 42.6 [15.7-74.8] |

^1^ Column percentages.

* Empty cell: no participants within this group of e-cigarette users fell into this subgroup.

##### Table S8. Sociodemographic and smoking profile of adult (≥16y) e-cigarette users in Great Britain in 2023: current smokers (*n*=820)

|  | **All**  **e-cigarette users**  **%^1^ (95%CI)** | **Disposable**  **e-cigarette users**  **%^1^ (95%CI)** | **Refillable**  **e-cigarette users**  **%^1^ (95%CI)** | **Pod**  **e-cigarette users**  **%^1^ (95%CI)** |
| --- | --- | --- | --- | --- |
| *Unweighted N* | *820* | *392* | *305* | *93* |
| Country of residence |  |  |  |  |
| England | 90.5 [88.8–92.0] | 91.6 [89.2–93.5] | 88.2 [84.9–90.8] | 90.1 [84.1–94.1] |
| Wales | 3.1 [2.3–4.1] | 2.2 [1.5–3.4] | 4.9 [3.2–7.3] | 2.3 [0.8–6.6] |
| Scotland | 6.4 [5.2–7.8] | 6.2 [4.6–8.3] | 6.9 [5.1–9.4] | 7.5 [4.3–12.9] |
| Age (years) |  |  |  |  |
| 16-24 | 30.0 [26.4–33.7] | 37.6 [32.2–43.3] | 22.3 [17.3–28.3] | 22.8 [14.3–34.5] |
| 25-34 | 24.9 [21.6–28.5] | 25.6 [21.0–31.0] | 24.5 [19.2–30.7] | 21.3 [13.4–32.2] |
| 35-44 | 17.0 [14.3–20.1] | 16.0 [12.2–20.7] | 18.1 [13.8–23.5] | 17.1 [10.3–27.1] |
| 45-54 | 11.8 [9.7–14.3] | 9.2 [6.6–12.7] | 13.7 [10.1–18.3] | 12.4 [7.0–21.0] |
| 55-64 | 11.8 [9.7–14.3] | 8.9 [6.3–12.3] | 14.8 [11.1–19.5] | 18.1 [10.9–28.6] |
| ≥65 | 4.5 [3.3–6.2] | 2.7 [1.5–4.8] | 6.6 [4.2–10.3] | 8.2 [4.0–16.1] |
| Gender |  |  |  |  |
| Men | 52.8 [48.9–56.6] | 48.8 [43.2–54.5] | 58.4 [52.0–64.5] | 51.0 [39.6–62.2] |
| Women | 44.9 [41.0-48.8] | 49.3 [43.7-55.0] | 38.4 [32.4–44.8] | 48.1 [36.9–59.5] |
| Other | 2.4 [1.6-3.6] | 1.9 [1.0-3.5] | 3.2 [1.8–5.7] | 1.0 [0.1–6.9] |
| Social grade C2DE (less advantaged) | 55.0 [51.2–58.7] | 55.2 [49.7–60.6] | 55.8 [49.5–61.8] | 50.7 [39.5–61.8] |
| Post-16 qualifications | 64.2 [60.3–67.9] | 65.7 [60.1–70.9] | 63.1 [56.6–69.1] | 55.3 [43.8–66.4] |
| Ethnic minority | 12.6 [10.4–15.2] | 12.6 [9.7–16.3] | 12.9 [9.3–17.7] | 14.2 [7.9–24.3] |
| ≥1 children in the household | 31.7 [28.1–35.5] | 35.2 [29.8–40.9] | 29.0 [23.4–35.3] | 23.6 [14.9–35.1] |
| History of ≥1 diagnosed mental health conditions | 53.1 [48.6–57.6] | 53.3 [46.7–59.8] | 53.1 [45.6–60.5] | 44.4 [31.8–57.7] |

^1^ Column percentages.

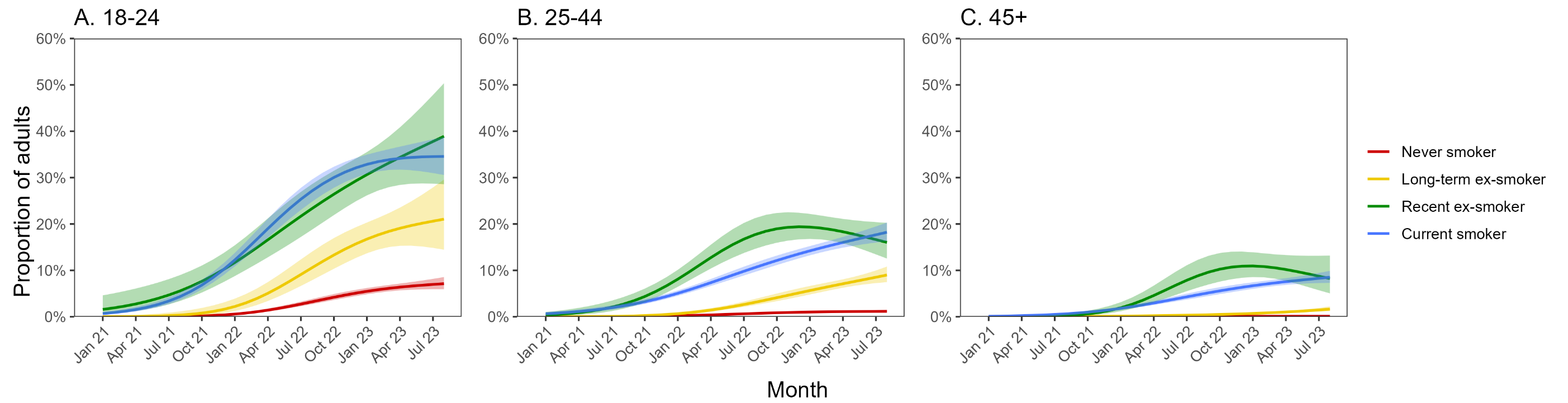

##### Figure S1. Trends in disposable e-cigarette use among adults (≥18y) in Great Britain, January 2021 to August 2023, by smoking status and age. Panels show smoking status-specific trends within adults aged (A) 18-24, (B) 25-44, and (C) ≥45 years. Lines represent modelled weighted prevalence by monthly survey wave, modelled non-linearly using restricted cubic splines (three knots). Shaded bands represent standard errors.

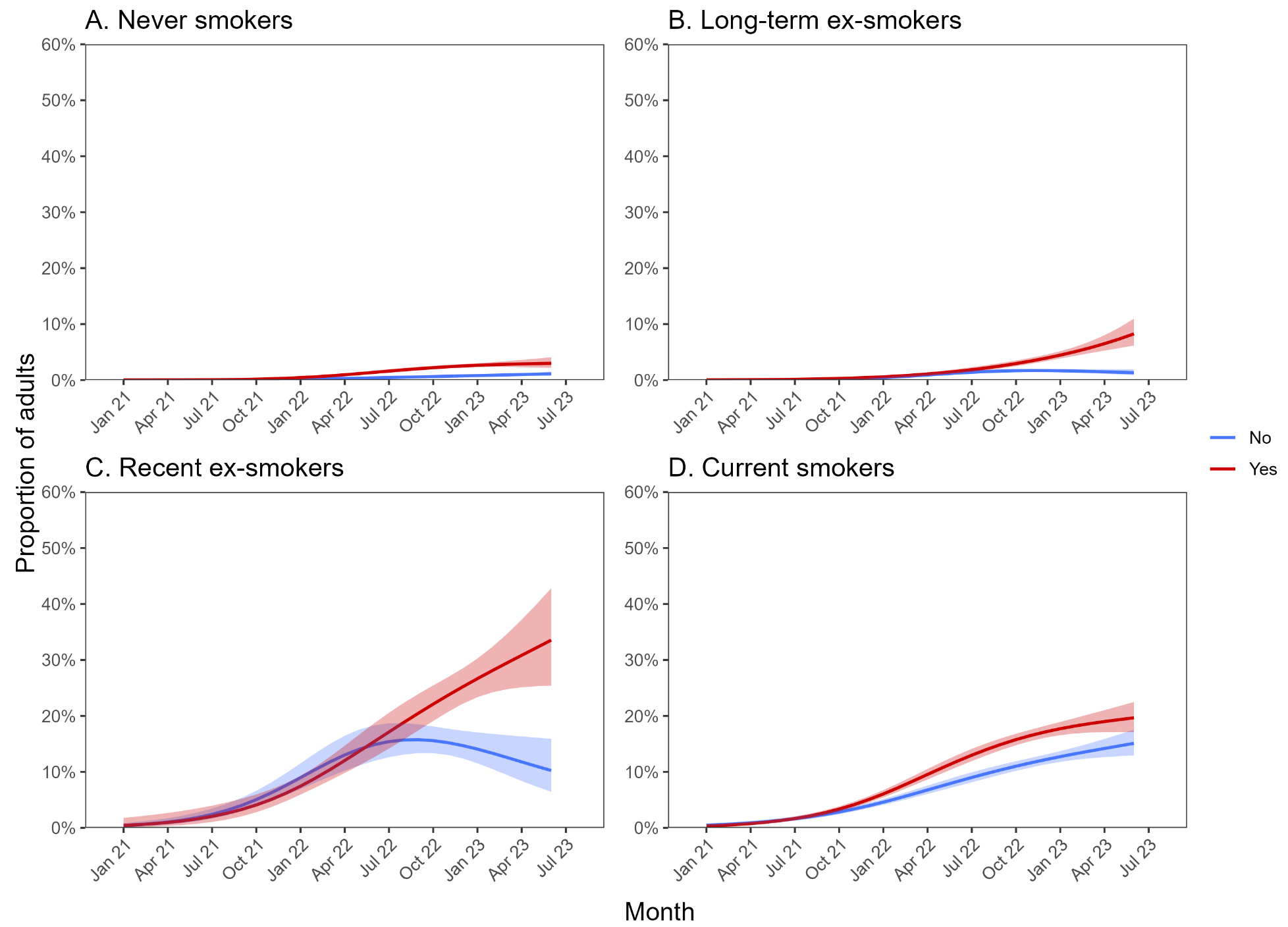

##### Figure S2. Trends in disposable e-cigarette use among adults (≥18y) in Great Britain, January 2021 to August 2023, by history of mental health conditions and smoking status. Panels show mental health-specific trends within adults who (A) have never regularly smoked, (B) stopped smoking ≥1 year ago, (C) stopped smoking in the past year, and (D) currently smokes. Lines represent modelled weighted prevalence by monthly survey wave, modelled non-linearly using restricted cubic splines (three knots). Shaded bands represent standard errors.

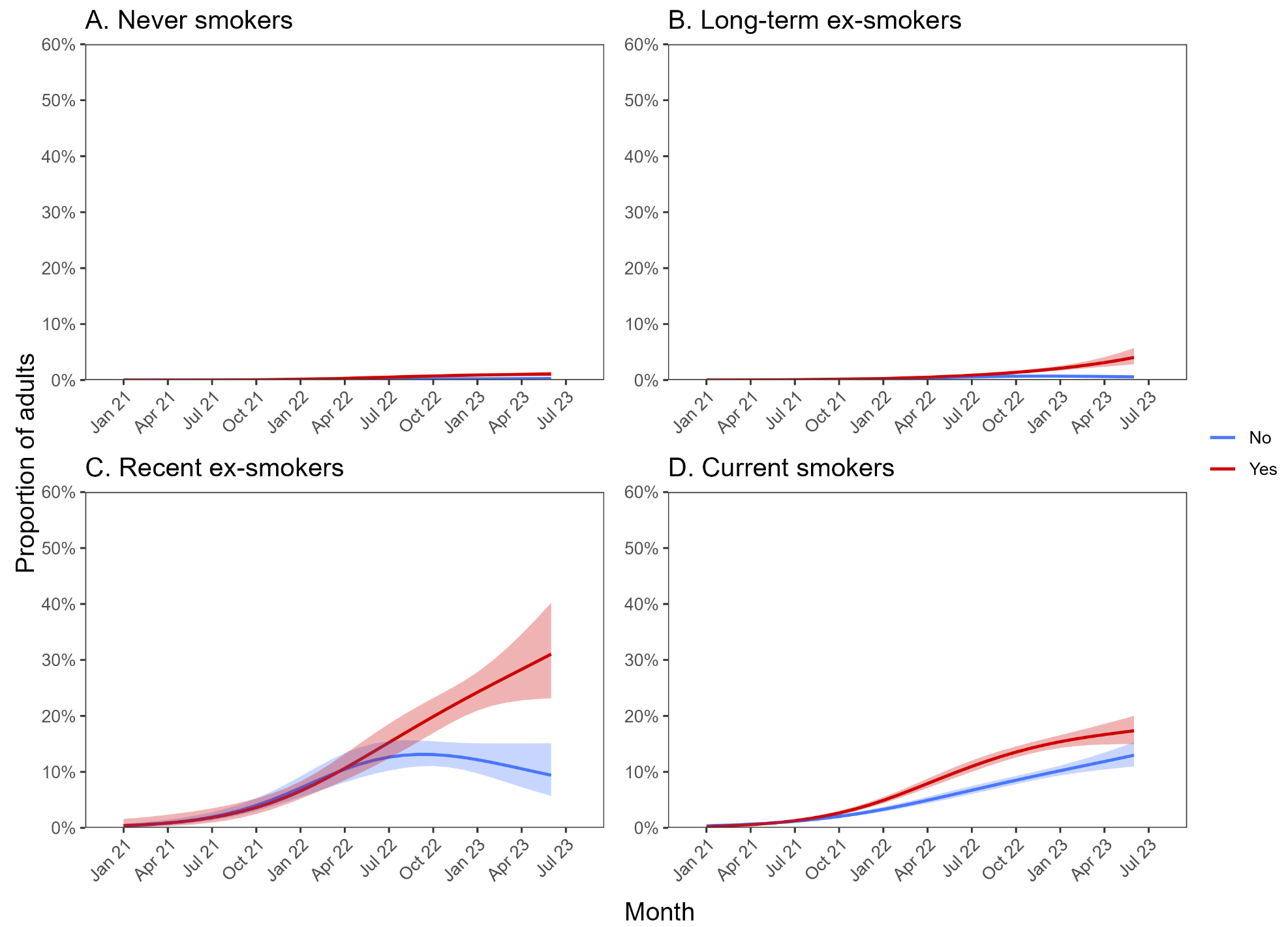

##### Figure S3. Age-adjusted trends in disposable e-cigarette use among adults (≥18y) in Great Britain, January 2021 to August 2023, by history of mental health conditions and smoking status. Panels show mental health-specific trends within adults who (A) have never regularly smoked, (B) stopped smoking ≥1 year ago, (C) stopped smoking in the past year, and (D) currently smokes. Lines represent modelled weighted prevalence by monthly survey wave, modelled non-linearly using restricted cubic splines (three knots). Shaded bands represent standard errors.
